# BSO-AD: An Ontology for Representing and Harmonizing Behavioral Social Knowledge in ADRD

**DOI:** 10.64898/2026.03.30.26349756

**Authors:** Haifang Li, Yue Yu, Avanti Bhandarkar, Rakesh Kumar, Isaac H. Clark, Yutong Hu, Weiguo Cao, Na Zhao, Fang Li, Cui Tao

**Affiliations:** Department of Artificial Intelligence and Informatics, Mayo Clinic, Jacksonville, FL 32224, USA; Department of Computer Science, Emory University, Atlanta, GA 30322, USA; Department of Neuroscience, Mayo Clinic, Jacksonville, FL 32224, USA

## Abstract

Behavioral and social factors (BSFs) substantially influence the risk, onset, and progression of Alzheimer’s disease and related dementias (ADRD), yet BSF-related knowledge remains scattered across heterogeneous sources, posing critical challenges for data harmonization and evidence synthesis. To address this gap, we present the **B**ehavioral **S**ocial Data and Knowledge **O**ntology for **AD**RD (BSO-AD), a FAIR-compliant semantic resource for representing and harmonizing BSFs and ADRD-related knowledge. BSO-AD was developed following established ontology design principles with reuse of existing ontologies and controlled terminologies, including the Social Determinants of Health Ontology, Drug Repurposing-Oriented Alzheimer’s Disease Ontology, AD-Onto, ICD-9-CM, and ICD-10-CM. Relationships between BSFs and ADRD were derived through literature mining. BSO-AD contains 1,690 classes, 152 object properties, 49 data properties, and 40 annotation properties. Ontology evaluation through Hootation-based domain expert review and a scalable LLM-assisted ontology assessment framework demonstrated high domain coverage and strong semantic coherence. BSO-AD provides a semantic foundation for data harmonization and knowledge integration in BSF-related ADRD research, while the LLM-assisted evaluation framework supports scalable and automated ontology assessment.

## INTRODUCTION

Alzheimer’s disease and related dementias (ADRD) constitute a major and escalating public health crisis^1^. In 2025, an estimated 7.2 million Americans aged 65 and older, about 1 in 9 people in this age group, are living with Alzheimer’s dementia (AD). This number is projected to nearly double to 13.8 million by 2060^2^. Along with increasing prevalence and mortality, ADRD imposes tremendous caregiving and financial burdens on patients, families, and healthcare systems^3^. Accumulating evidence demonstrates that behavioral and social factors (BSFs) play a pivotal role in shaping the risk, progression, and outcomes of ADRD across populations^4^. BSFs encompass behavioral and social determinants of health (e.g., physical activity, smoking, and social isolation) that influence neurocognitive resilience and aging trajectories^5^. Importantly, BSFs contribute to observed disparities in ADRD incidence, diagnosis, and care by reflecting differences in healthcare access and socioeconomic conditions^6^. Integrating BSF knowledge into ADRD prevention and intervention frameworks is critical for informing the development of targeted health policies and enhancing the quality of life for affected individuals and families^7,8^.

However, systematic integration of BSF knowledge into ADRD research remains a major challenge due to the heterogeneity of data sources and variability in their representation. In structured data settings, BSF information, derived from surveys, electronic health records (EHRs), and mobile apps, is often encoded using inconsistent standards and value sets. Unstructured sources, including clinical narratives and scientific literature, contain rich contextual information but pose substantial challenges for extraction, normalization, and computational analysis. Moreover, integrating BSF and ADRD data, both structured and unstructured, requires cross-disciplinary alignment, as differences across domain-specific conceptual systems further impede effective aggregation. These challenges necessitate a robust, interoperable ontological infrastructure to harmonize heterogeneous data sources and enable standardized, AI-driven analysis of BSFs in ADRD.

Several AD-specific ontologies have been developed, each with distinct focuses, structures, and applications. For example, the Alzheimer’s Disease Ontology (ADO) provides a fine-grained hierarchy capturing biological and clinical dimensions^9^ while the Drug Repurposing-Oriented Alzheimer’s Disease Ontology (DROADO)^10^ integrates AD-related drugs, genes, targets, and pathways to support computational drug repurposing. Parallel efforts to standardize BSFs using ontologies have emerged. The Ontology of Medically Related Social Entities (OMRSE) captures health-related societal entities^11^, while the Semantic Mining of Activity, Social, and Health data (SMASH) ontology models biomarkers, social activities, and physical activities associated with sustained weight loss^12^. The Social Determinants of Health Ontology (SDoHO) further defines core SDoH factors and their relationships within a structured, measurable framework^13^. Despite these advances, existing ontologies remain largely siloed, focusing either on the biological-clinical aspect of ADRD or on BSFs in isolation. This fragmentation represents a critical unmet need for an integrated ontology that systematically unifies the representation of BSFs and ADRD within a coherent, forward-looking framework.

To address this need, we developed the **B**ehavioral **S**ocial Data and Knowledge **O**ntology for **AD**RD (BSO-AD) to harmonize the conceptual systems of these two domains by reusing and extending established ontologies and standards. To capture the multifaceted relationships between BSFs and ADRD, we modeled both direct and mechanistic associations. Direct associations were synthesized through literature review, whereas mechanistic associations were formalized via intermediate biological entities, including genes, pathways, and pathological processes, realizing multi-level, biologically grounded representations. BSO-AD was assessed through expert review of its semantics, using the ontology evaluation tool Hootation^14^. Furthermore, we developed a large language model (LLM)-assisted, domain-informed evaluation pipeline for automated ontology assessment. By bridging traditional siloed behavioral-social, biological, and medical domains within a unified semantic framework, BSO-AD provides a robust knowledge infrastructure for interoperable data integration and scalable AI-driven applications, ultimately advancing evidence-based strategies for ADRD prevention and intervention.

The main contributions of this study are as follows:

- To the best of our knowledge, BSO-AD represents the first ontology that systematically formalizes BSFs in the context of ADRD.
- We designed a multi-layer relationship framework to capture both direct associations between BSFs and ADRD and indirect relationships mediated by underlying biological mechanisms and interactions.
- We developed an LLM-assisted, data-driven framework to enable scalable, automated ontology evaluation.

## METHODS

### Ontology Development

BSO-AD was represented in the Web Ontology Language (OWL2)^15^. We used ROBOT^16^ for modular assembly and template-based ontology format conversion, followed by manual curation in Protégé 5.6.4^17^. The BSO-AD development framework was illustrated in **Figure 1**.

**Figure 1.**
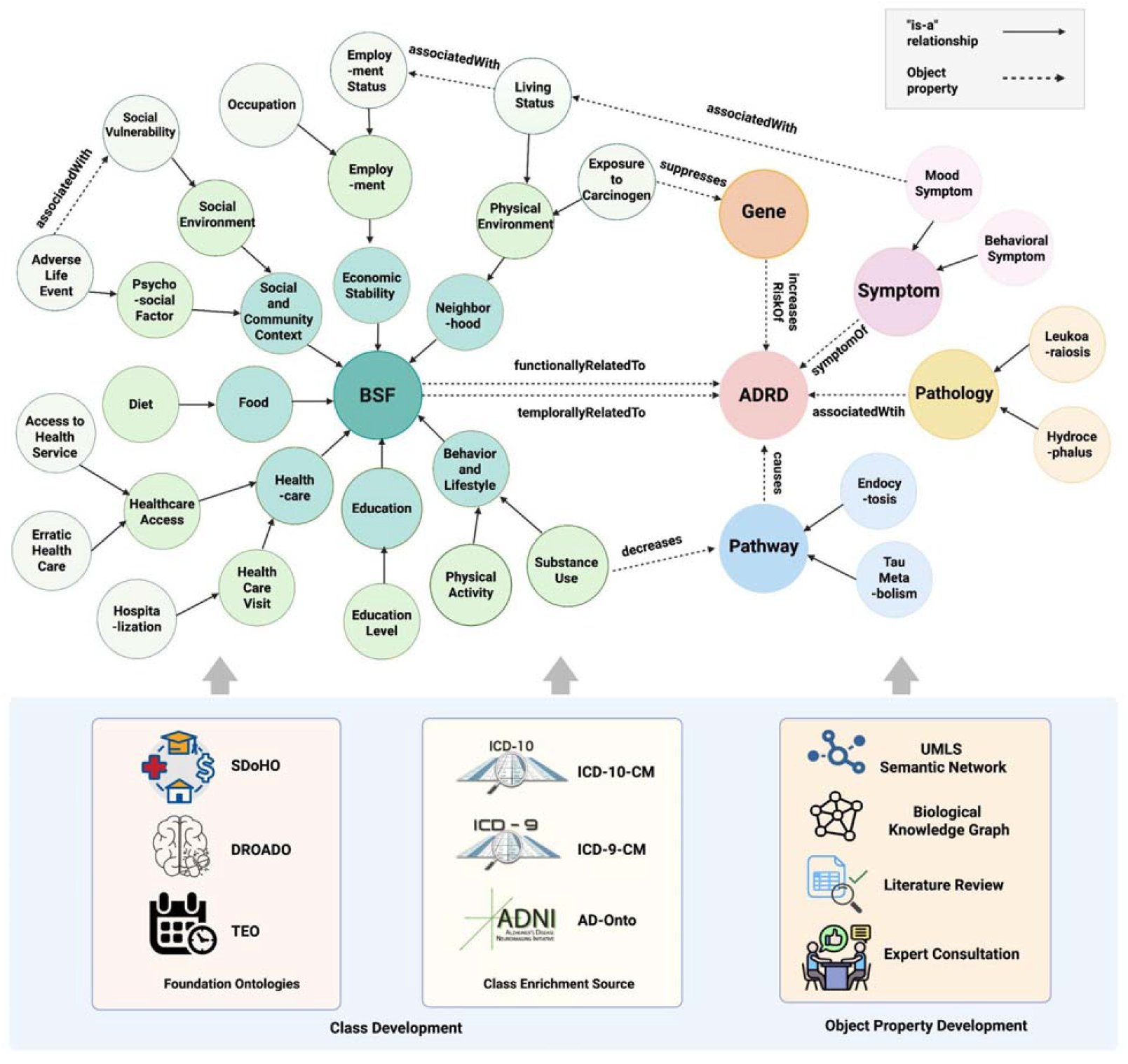
BSO-AD Development Framework

Development framework of the BSO-AD. The ontology development process consists of class development and object property development. Class development included ontology reuse from foundation ontologies, including SDoHO, DROADO, and TEO, as well as class enrichment using ICD-10-CM, ICD-9-CM, and AD-Onto. Object property development was guided by the UMLS semantic network, relevant biomedical knowledge graphs, literature review, and domain expert consultation. BSO-AD primarily comprises BSF categories, ADRD-related classes, and the hierarchical (is-a) and semantic (object property) relationships connecting these classes.

Abbreviations: ADNI: Alzheimer’s Disease Neuroimaging Initiative; ADRD: Alzheimer’s Disease and Related Dementia; BSF: Behavioral and social factor; DROADO: Drug Repurposing-Oriented Alzheimer’s Disease Ontology; ICD-9-CM: International Classification of Diseases, Ninth Revision, Clinical Modification; ICD-10-CM: International Classification of Diseases, Tenth Revision, Clinical Modification; SDoHO: Social Determinants of Health Ontology; TEO: Time Event Ontology.

#### Principles

The development of BSO-AD followed the best practices in biomedical ontology engineering, including the Open Biological and Biomedical Ontologies (OBO) Foundry principles^18^ and the FAIR principles (Findable, Accessible, Interoperable, and Reusable)^19^. Collectively, these frameworks emphasize openness, community collaboration, interoperability, and reusability. A central design principle of BSO-AD is ontology reuse with modular extension, prioritizing reuse and alignment with established ontologies and controlled vocabularies. This strategy enhances semantic interoperability and minimizes redundancy^20,21^. New classes and properties are introduced only when gaps are identified, and no suitable ones are available in established ontologies or standards.

#### Class Design

The construction of the BSO-AD class hierarchy followed a combination of ontology reuse^20,21^, top-down^22^, and bottom-up^22^ approaches. The top-down approach begins with broad domain concepts that are progressively specialized, while the bottom-up approach identifies specific classes from data or literature and groups them into higher-level categories^22^.

Three existing ontologies were incorporated into BSO-AD based on domain relevance and modeling scope: SDoHO^13^, DROADO^10^, and the Time Event Ontology (TEO)^23^. SDoHO was reused to represent the BSFs, as it provides a standardized and data-driven framework for modeling SDoH and their interrelationships^24^. DROADO^10^ was integrated to capture ADRD-related medical and biological knowledge, including disease and molecular mechanisms. In addition, TEO^23^ was included to formally represent the temporal dimension within the ADRD context.

Although SDoHO provides a comprehensive conceptual framework for modeling BSFs, it has not incorporated the relevant concepts from standard clinical coding systems such as the International Classification of Diseases (ICD)-10-Clinical Modification (CM) for EHR interoperability. To address this limitation, we integrated SDoH-related ICD-10-CM Z55–Z65 codes^25^, which document patients’ health hazards related to socioeconomic, occupational, and psychosocial circumstances^26^. Following the bottom-up approach^22^, these concepts were either merged with existing classes or created as new subclasses under appropriate branches of the BSF hierarchy. Notably, ICD includes residual categories containing terms such as “other” or “unspecified” (e.g., “Other problems related to education and literacy” and “Unemployment, unspecified”). To align with the principle of clear and unambiguous naming advocated by the OBO Foundry^18^, we mapped these concepts to their parent classes, using annotation property *skos:narrower*. For example, “Z56.0 Unemployment, unspecified” was mapped to the class “Unemployment”, with annotation property: “skos:narrower: Z56.0”, accompanied by sub-annotations including “dc:source: ICD-10-CM” and “Description: Unemployment, unspecified”. Details of these concepts and their hierarchical placement within the ontology (i.e., parent classes) are provided in **Supplementary 1**.

While DROADO captures molecular and pharmacological knowledge relevant to ADRD, it provides limited coverage of neuropsychological assessments, which are critical for ADRD diagnosis and monitoring. Therefore, we incorporated branches from AD-Onto^27^, a computational ontology focusing on the neuropsychological tests derived from the Alzheimer’s Disease Neuroimaging Initiative (ADNI) data collection. Specifically, we reused the “Examination” and “StandardizedAssessmentItem” branches, which represent various clinical evaluation types (e.g., neuropsychological and vascular risk assessments) and cognitive assessment components (e.g., MMSE subitems such as orientation, recall, and registration). Additionally, following the bottom-up approach^22^, ADRD-related concepts from ICD-9-CM and ICD-10-CM (**Supplementary 2, Table S1**) were integrated to expand coverage of ADRD concepts.

#### Property Design

##### Object property

In OWL ontologies, object properties describe binary semantic relationships between two individuals (or instances)^28^, thereby encoding knowledge that extends beyond taxonomic hierarchies. We reused the necessary object properties from the source ontologies (SDoHO, DROADO and TEO), as well as those associated with the reused “Examination” and “StandardizedAssessmentItem” branches from AD-Onto, to preserve the important semantic relationships among the domain concepts.

Systematic representation of the potential semantic relationships between BSFs and ADRD was a high priority in designing BSO-AD object properties. We conducted a targeted PubMed literature search (query detailed in **Supplementary 2, Table S2**) and manually reviewed the retrieved abstracts (n = 216) to summarize potential relationships. These relationships were subsequently aligned with relation types defined in the Unified Medical Language System (UMLS) Semantic Network^29^ to ensure semantic standardization and interoperability.

Furthermore, beyond the direct associations, a substantial body of research has reported biological mechanisms by which BSFs affect ADRD^30–32^. To uncover the mechanistic pathways and support multi-level knowledge representations, we incorporated three key biological entity types, i.e., genes, pathways, and pathologies, as intermediate nodes. Relationships among BSFs, three intermediate nodes, and ADRD were modeled based on evidence derived from the literature and refined through consultation with domain experts.

##### Annotation and Data Property

Annotation properties provide additional information for describing and labeling concepts in an ontology^33^. Several standardized IDs, such as UMLS Concept Unique Identifiers (CUIs), ICD-10-CM codes, and ICD-9-CM codes, were added as annotation properties to facilitate semantic interoperability with external terminologies and clinical coding systems. In addition, annotation properties from source ontologies, including comments, definitions, and provenance information, were also inherited.

Data properties link individuals to literal values (e.g., strings, numbers)^15^. For BSO-AD, we primarily reused the data properties from source ontologies, including SDoHO, DROADO, AD-Onto, and TEO.

### Ontology Evaluation

#### Hootation-Based Expert Review

First, we evaluated the ontology semantics through expert review. The ontology was transformed into natural language sentences using the ontology evaluation tool Hootation^14^. Two domain experts independently assessed each sentence to determine whether the corresponding ontology assertion (i.e., every child class is a type of its parent class) was semantically valid. Two agreement metrics were calculated: (1) Inter-evaluator agreement, defined as the proportion of statements for which both evaluators assigned the same label; and (2) Rational agreement, defined as the proportion of statements jointly judged as rational (i.e., the number of statements labeled as rational by both evaluators divided by the total number of statements)^13^. Disagreements were resolved through discussion to achieve consensus.

#### LLM-Assisted, Data-Driven Framework for Automated Evaluation

To enable automated and scalable evaluation, we developed a domain-informed, LLM-assisted framework to assess BSO-AD. By integrating multiple open-source LLMs (Llama4^34^, Qwen3^35^, and MedGemma^36^) within a unified pipeline for systematic literature selection and entity extraction, our framework evaluated representational breadth and conceptual coherence within the BSF-ADRD domain, grounded in evidence derived from the literature (**Figure 2**).

**Figure 2.**
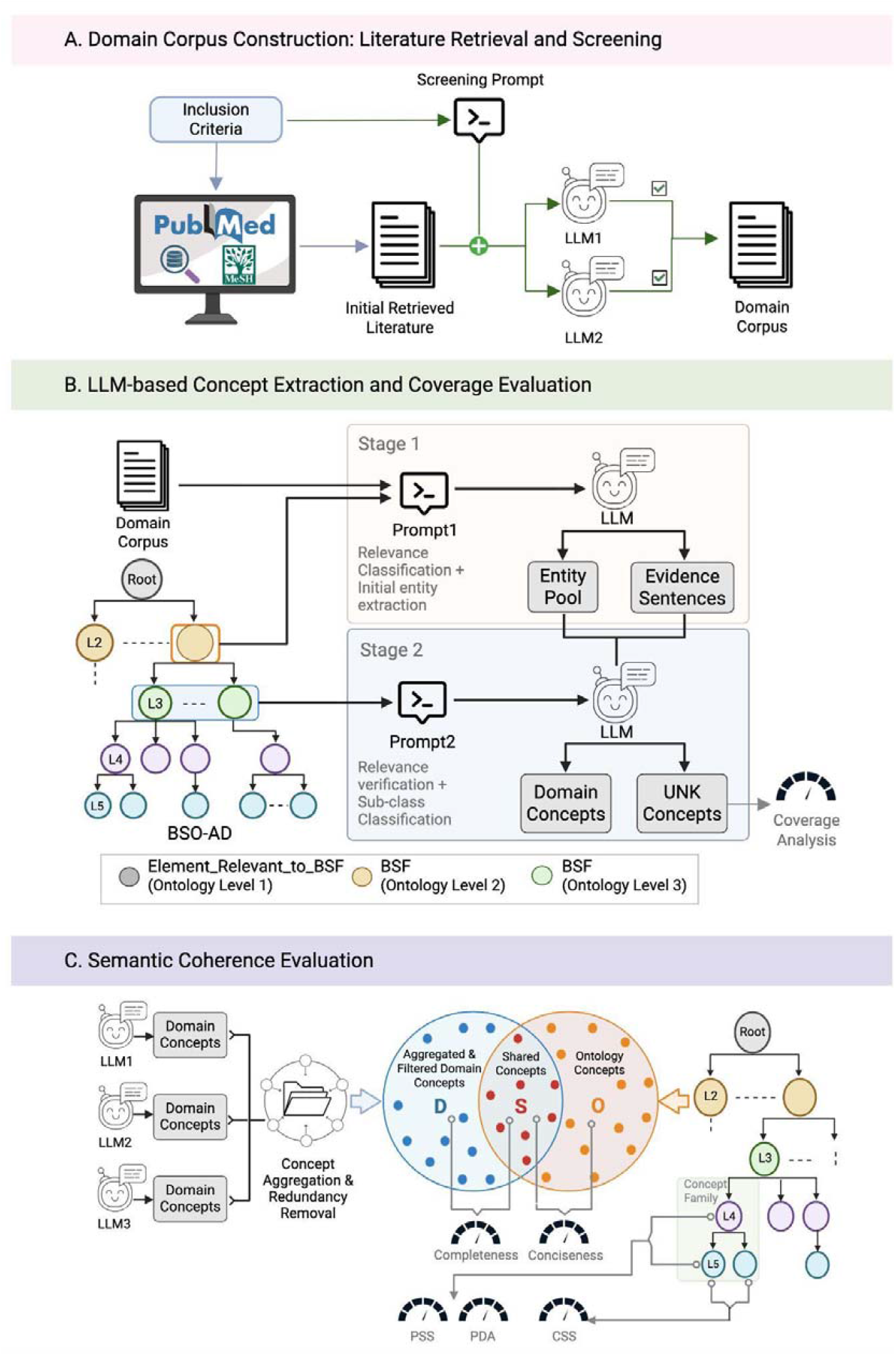
LLM-assisted, Data-Driven Framework for Automated Ontology Evaluation. (A) Construction of the targeted domain corpus. (B) LLM-assisted and ontology-guided concept extraction for BSF category-level coverage evaluation. (C) LLM-annotated concept aggregation and embedding-based semantic coherence evaluation, including Completeness, Conciseness, CSS, PSS, and PDA. Abbreviations: BSF: Behavioral and social factor; CSS: Child Similarity Score; PDA: Parent-Child Difference Agreement; PSS: Parent-Child Similarity Score.

##### Domain Literature Corpus Construction

To create a representative domain corpus, we retrieved PubMed abstracts on ADRD and BSFs using Medical Subject Headings (MeSH)^37^ terms and keyword-based queries (**Supplementary 2, 2.1.1**). Two general-purpose LLMs (Llama4 and Qwen3) independently screened each article’s title and abstract against pre-defined inclusion criteria (**Supplementary 2, 2.1.2**). Only articles with consensus by the two LLMs were retained to ensure relevance and coverage (**Figure 2A**).

##### Concept Extraction and Category Coverage Evaluation

To assess how effectively BSO-AD captures concepts from the literature, we designed a two-stage, ontology-guided framework for concept extraction, classification, and coverage evaluation (**Figure 2B**). In the first stage, three LLMs, including one medical-specific model (MedGemma) and two general-purpose models (Llama4 and Qwen3), independently screened the title and abstract of selected articles (from Step 1 Domain Literature Corpus Construction) to extract ADRD-related BSF entities. In the second stage, each model assigned the extracted entities to BSF subcategories using ontology-informed definitions and few-shot prompting. Entities that could not be classified into any BSF subcategories were labeled as “Undefined” (UNK). The prompts for this process are provided in **Supplementary 2, 2.2**.

Category coverage rate was defined as the proportion of extracted entities that were successfully assigned or categorized to ontology classes (i.e., not labeled as Undefined). This metric was computed based on entities identified and classified by MedGemma, given its domain specialization^1^.

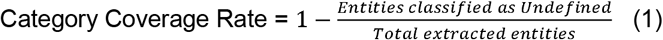

##### Semantic Coherence Evaluation

To assess BSO-AD’s semantic adequacy beyond explicit class mappings, we adopted and extended the ontology evaluation framework proposed by Zaitoun et al.^38^ (**Figure 2C**). Unlike the original framework that relied on a pre-trained named entity recognition (NER) model for concept extraction, we utilized the ontology-guided, LLM-based extraction and classification framework described above to enable broader and more flexible capture of domain concepts across clinical and socio-behavioral dimensions. Evaluations were performed across seven BSF sub-categories using ClinicalBERT^39^.

To ensure comprehensive capture of domain concepts, the entity extraction and classification process was conducted by three LLMs individually. Concepts identified as BSF-relevant by any LLM were aggregated and deduplicated based on cosine similarity in the embedding space, with FAISS^40^ used for efficient nearest-neighbor search.

Entities paired with similarity ≥ 0.90 were treated as semantically equivalent and merged, yielding the domain concept set. A shared concept was defined as an ontology concept having at least one nearest (cosine similarity ≥ 0.75) domain concept in the embedding space, indicating meaningful semantic overlap despite lexical variations.

#### Completeness and Conciseness

Completeness is defined as the proportion of domain concepts captured by the ontology, reflecting coverage of domain knowledge derived from the literature, whereas conciseness is defined as the proportion of ontology classes that correspond to domain concepts, indicating the extent to which the ontology concepts are grounded in domain evidence while avoiding redundancy. These two metrics are defined as follows^38^:

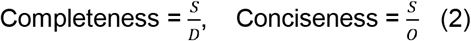

Where O denotes the number of concepts in the ontology, D denotes the number of domain concepts extracted from literature using LLMs, and S denotes the number of shared concepts between them.

#### Correctness and Consistency

Semantic correctness and ontology hierarchy consistency were evaluated following prior ontology evaluation metrics, including the Child Similarity Score^38^ for semantic correctness , and the Parent-Child Similarity Score^38^ and Parent-Child Difference Agreement^38^ for ontology hierarchy consistency. These metrics were computed based on concept families (CFs), each consisting of a parent concept and its direct child concepts. In all metrics, similarity denotes cosine similarity between ClinicalBERT-based concept embeddings.

Child Similarity Score (CSS) measures the average pairwise cosine similarity among sibling concepts within a CF^38^.

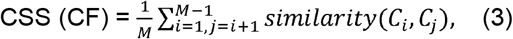

where *M* denotes the number of child concepts in the CF, and *C*_*i*_ and *C*_*j*_ denote sibling (child) concepts.

Parent-Child Similarity Score (PSS) measures the average cosine similarity between a parent concept and its direct child concepts^38^.

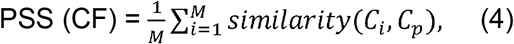

where *C*_*p*_ denotes the parent concept, *C*_*i*_ denotes a child concept, and *M* is the number of child concepts.

Parent-Child Difference Agreement (PDA) evaluates the consistency of cosine similarity between child concepts and their parent concept^38^.

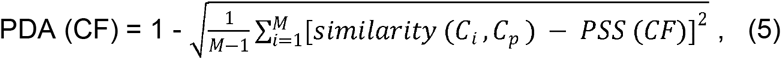

where *C*_*p*_ denotes the parent concept, *C*_*i*_ denotes a child concept, *M* denotes the number of child concepts, and PSS(CF) denotes the Parent-Child Similarity Score of the CF.

For ease of reference, all abbreviations are listed in **Supplementary 2, Table S5**.

## RESULTS

### Ontology

BSO-AD provides a comprehensive framework for representing BSF knowledge in the context of ADRD, with a well-organized class hierarchy and clearly defined object, data, and annotation properties. The current version of BSO-AD comprises 1,690 classes, 152 object properties, 49 data properties, and 40 annotation properties with 2,535 logical axioms and 2,001 declaration axioms.

#### Classes

The current BSO-AD includes 14 root classes that represent BSFs and ADRD-related data and knowledge (**Figure 3**), including 3 from SDoHO (i.e., “Element_Relevant_to_Bahavioral_Social_Factor”, “Measure_and_Index_and_Score”, and “Person”), 9 from DROADO (e.g., “Disease” and “Gene”), one from TEO (“Element_relevevant_to_Time_Event”), and one from AD-Onto (“Element_Relvant_to_Examination”).

**Figure 3.**
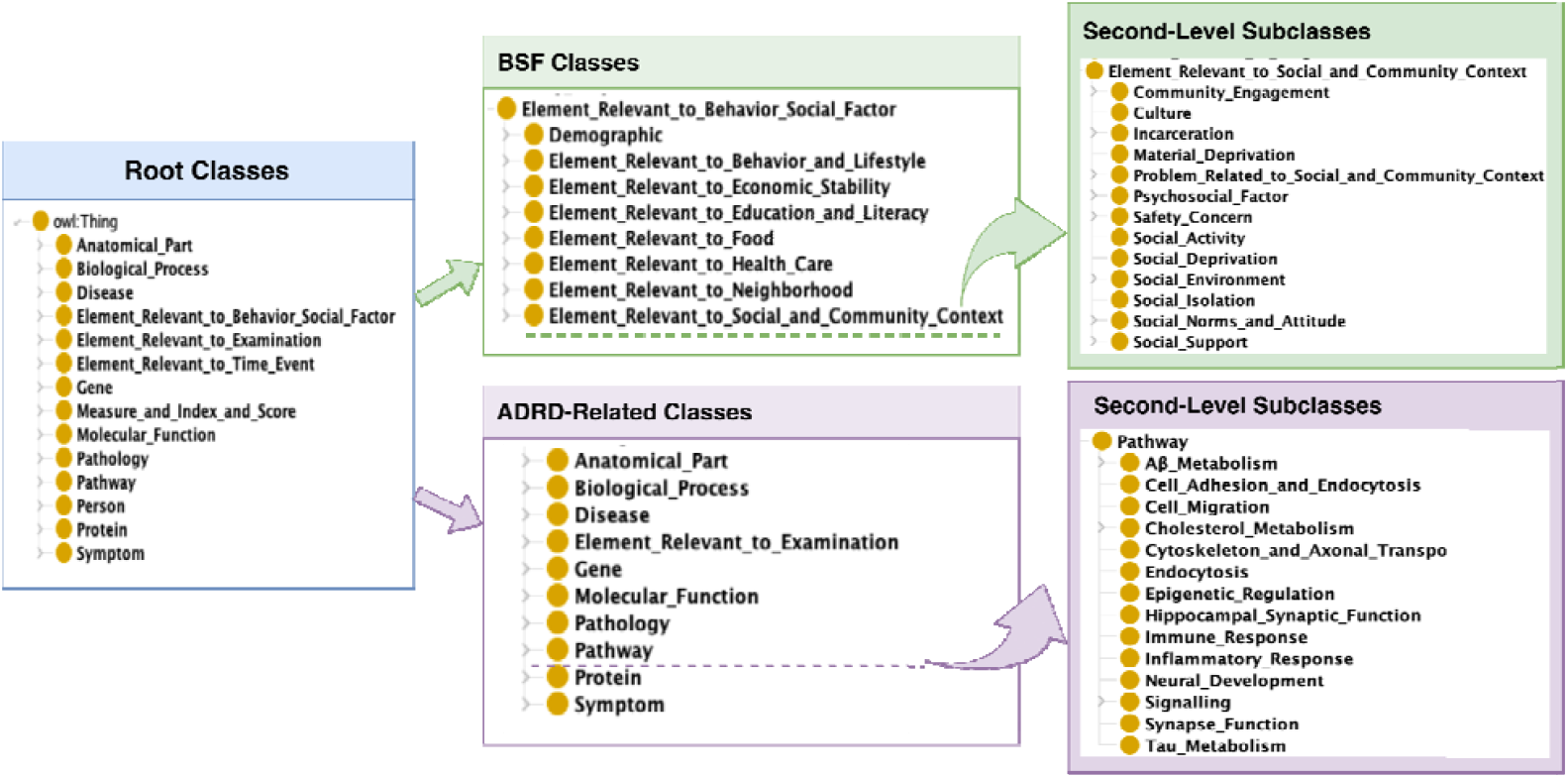
The Hierarchy of BSO-AD

Within “Element_Relevant_to_Behavioral_Social_Factor”, 8 major branches represent the core BSFs, such as “Element_Relevant_to_Food” and “Element_Relevant_to_Neighborhood”, which were imported from SDoHO. In addition, a total of 142 relevant concepts are imported from ICD-10-CM codes Z55–Z65 (**Supplementary 1**), and subsequently mapped and merged into the behavioral-social hierarchies.

ADRD-related knowledge is represented by 10 branches, of which 9 were imported from DROADO, and “Element_Relvant_to_Examination” from AD-Onto. Furthermore, 21 ADRD-related concepts (**Supplementary 2, Table S1**) from ICD-9-CM and ICD-10-CM were included. Finally, “Element_Relvant_to_Time_Event” branch captures temporal dimensions, all of which were adopted from the TEO ontology.

#### Properties

The BSO-AD defines 152 object properties, 49 data properties, and 40 annotation properties, which together formalize the semantic relationships and descriptive characteristics within and across BSFs and ADRD domains.

We defined a multi-layer relational framework to capture both the direct and indirect associations of BSFs with ADRD. Specifically, 26 object properties are defined to represent direct associations, which are organized into a multi-level hierarchy grounded in the UMLS Semantic Network (**Table 1**). Under the top-level “associatedWith” category, there are two major types of semantic relationships: “functionallyRelatedTo” and “temporallyRelatedTo”. The “functionallyRelatedTo” captures function-based associations and further expands to “affects” and “predicts” subrelations. The “affects” is specialized into “affectsRiskOf”, “affectsDiseaseCourse”, and “affectsDiseaseBurden” to model the effects of BSFs on disease risk, progression, and disease-related burden on individuals or populations, respectively. “temporallyRelatedTo” describes time-dependent associations, such as “associatedWithShorterTimetoEvent”, supporting representing temporal dynamics (e.g., earlier onset or delayed progression). To facilitate downstream natural language processing tasks, property is enriched with a set of lexical variants and synonyms (e.g., “increasesRriskOf”: “raisesRiskOf”, “escalatesRiskOf”) by incorporating literature-derived expressions (**Supplementary 2, Table S3**).

**Table 1.** Object Properties between BSFs and ADRD under “associatedWith”.

| Relationship Name | Definition | Examples |
| --- | --- | --- |
| functionallyRelatedTo | An entity is associated with another entity through a functional, biological, clinical, or statistical relationship, encompassing effects, influences, or predictive associations. | Categorical Relationship |
| affects | A general relationship indicating that an entity influences another entity or process in any measurable or observable way, without specifying direction, magnitude, or mechanism. | Social health is increasingly recognised as an important factor *influencing* cognitive ageing and experiences of dementia. (PMID: 41936900) |
| affectsDiseaseCourse | An entity modifies the progression, severity, or temporal trajectory of a disease after its onset. | Social isolation has emerged as a significant social determinant that may *exacerbate* cognitive deterioration in older adults. (PMID: 41087920)<br>This study emphasizes the necessity of addressing S/SDOH to *mitigate* dementia risk among Black Americans. (PMID: 39612114) |
| complicates | Introduces additional clinical challenges, comorbidities, or variability that make disease management or progression more complex. |  |
| exacerbates | Worsens the severity or accelerates the progression of a disease. |  |
| mitigates | Reduces the severity or slows the progression of a disease. |  |
| affectsRiskOf | An entity modifies the probability that an individual will develop a disease or health outcome. | This study of older Latino adults demonstrated an association between reduced brain white matter integrity and contextually related socioenvironmental |
| decreasesRiskOf | Lowers the likelihood that an individual will develop a disease or outcome. |  |
| increasesRiskOf | Raises the likelihood that an individual will develop a disease or outcome. | experiences known to <u>increase risk of</u> MCI and Alzheimer's Disease. (PMID: 37718813) |
| contributesTo | An entity plays a partial or contributing role in the development of a disease or outcome, typically as one factor among multiple interacting causes. | It is well known that vascular factors and specific social determinants of health <u>contribute to</u> dementia risk and that the prevalence of these risk factors differs according to race and sex. (PMID: 37490246 ) |
| affectsDiseaseBurden | An entity modifies the overall impact of a disease on individuals or populations, including severity, disability, quality of life, or healthcare utilization. | These findings demonstrate the importance of involving those experiencing negative effects in R/S for <u>reducing the ADRD burden</u> for Black people in the US. (PMID: 39589019) |
| increasesBurdenFor | Increases the clinical, functional, or societal impact of a disease on individuals or populations. |  |
| reducesBurdenFor | Decreases the clinical, functional, or societal impact of a disease on individuals or populations. |  |
| affectsPopulationMetric | An entity modifies a measurable population-level health indicator. | Lower socioeconomic status (SES) and less education are associated with <u>a higher incidence of</u> ADRDs. (PMID: 34103175) |
| affectsIncidenceOf | An entity modifies the rate at which new cases of a disease occur in a population over a specified period. |  |
| decreasesIncidenceOf | Leads to a lower rate of new cases of a disease in a population over time. |  |
| increasesIncidenceOf | Leads to a higher rate of new cases of a disease in a population over time. |  |
| affectsPrevalenceOf | An entity modifies the proportion of individuals in a population who have a disease at a given time. |  |
| decreasesPrevalenceOf | Leads to a lower proportion of individuals with a disease in a population. |  |
| increasesPrevalenceOf | Leads to a higher proportion of individuals with a disease in a population. |  |
| negativelyAffects | An entity exerts an effect that worsens or increases adverse outcomes associated with a disease or health condition. | Acculturation may <u>benefit</u> cognition when SEP is low. ( PMID: 32440686 ) |
| positivelyAffects | An entity exerts an effect that improves or reduces adverse outcomes associated with a disease or health condition. |  |
| predicts | The presence or value of an entity can be used to estimate the likelihood or future occurrence of a disease or outcome, without implying causality. | The addition of known dementia risk factors in cluster analysis did not change the findings, suggesting that social determinants of health independently <u>predict</u> dementia risk. (PMID: 40110677) |
| temporallyRelatedTo | An entity is associated with the timing or temporal characteristics of a disease-related event or outcome, without specifying the direction or magnitude of the temporal effect. | Categorical Relationship |
| associatedWithLongerTimeToEvent | An entity is associated with an increased time interval to the occurrence of a specified disease-related event or outcome, such as delayed onset, slower progression, or prolonged survival time. | Older ETOA and higher amyloid are <u>associated with shorter time</u> from tau pathology onset (T+) to dementia. (PMID: 40859634) |
| associatedWithShorterTimeToEvent | An entity is associated with a reduced time interval to the occurrence of a specified disease-related event or outcome, such as earlier onset, faster progression, or shorter survival time. |  |

In parallel, we built indirect relationships to represent how BSFs may influence ADRD through intermediate biological nodes, including genes, pathways, and pathology (**Figure 4 and Supplementary 2, Table S4**), informed by heterogeneous biomedical frameworks such as Hetionet^41^. For example, a BSF may *modulate* a gene, the gene may *regulate* a pathway, the pathway may *perturb* a pathology, and the pathology may be *associatedWith* ADRD. By modeling these multi-level mechanistic linkages, we aimed to capture the underlying mechanisms through which BSFs contribute to the development and progression of ADRD.

**Figure 4.**
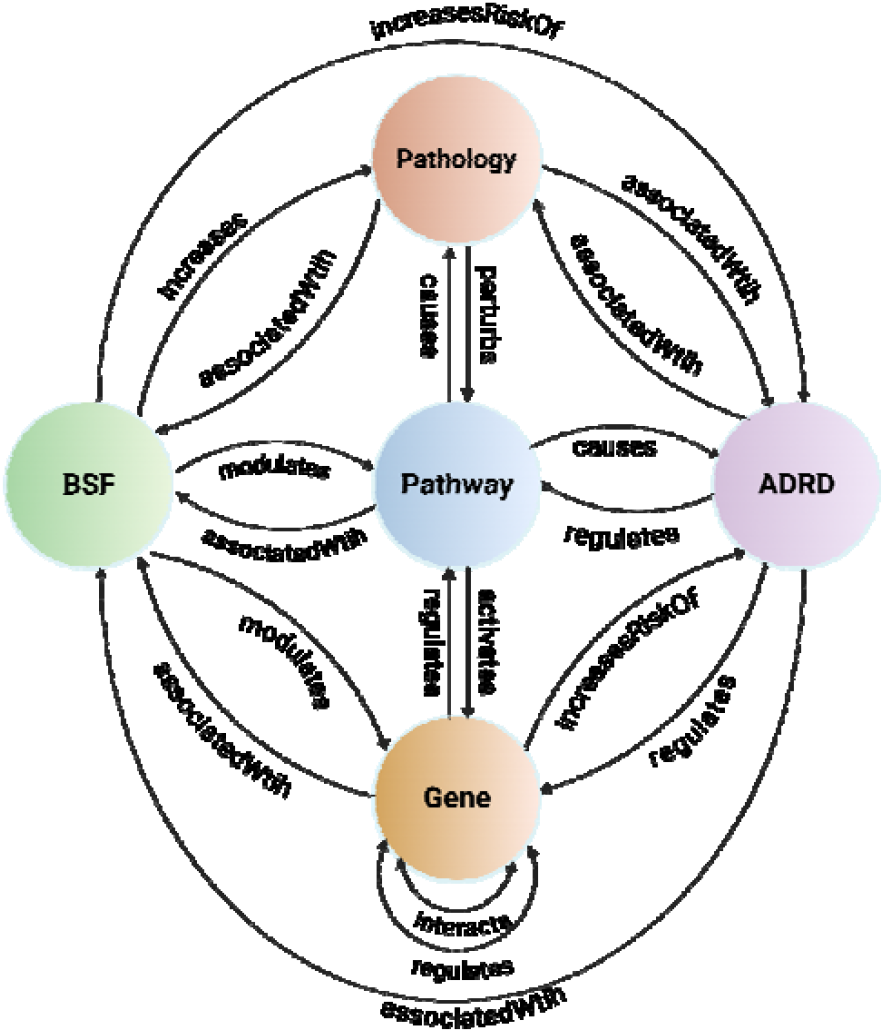
Indirect relationships connecting BSF and ADRD through intermediate biological nodes. Abbreviations: ADRD: Alzheimer’s Disease and Related Dementia; BSF: Behavioral and social factor.

The annotation properties include standard metadata elements such as comments, definitions, and provenance information. In addition, interoperability is strengthened by adding annotation properties such as *skos:exact, skos:narrower, dc:source* to connect CUIs and ICD-10-CM codes, facilitating integration with external terminologies and standards.

### Evaluation Results

#### Hootation-Based Expert Review

We evaluated the semantic quality of BSO-AD using Hootation, an ontology verbalization tool for converting OWL axioms into natural language text ^14^, with outputs independently assessed by two domain experts. The inter-evaluator agreement was 0.96, and disagreements were resolved through discussion, resulting in consensus across all classes. The rational agreement was 0.95, indicating that 95% of statements were judged semantically valid by both experts. Identified issues were addressed and incorporated into ontology refinement. For example, the statement “every Non-smoker is a type of Smoker” reflected a semantic conflict. Accordingly, we restructured “Non-smoker” and “Smoker” as sibling classes rather than a child-parent relationship. Similarly, for the problematic statement “every Absence_of_Family_Member is a type of Family_Support”, we introduced a new parent class, “Problem_Related_to_Family_Support”, to accommodate “Absence_of_Family_Member”, and positioned them in an appropriate hierarchy under the high-level class “Problem_Related_to_Social_and_Community_Context”.

#### LLM-based Automated Evaluation

As shown in **Table 2**, category coverage rates were uniformly high across all BSF domains (≥ 0.97), indicating that most literature-derived concepts were successfully classified into BSO-AD’s high-level classes. Detailed category-wise results are provided in **Supplementary 2, 2.3**. Completeness averaged 0.78, reflecting substantial alignment between ontology concepts and literature-derived domain concepts. Meanwhile, Conciseness averaged 0.81, suggesting that a large proportion of ontology classes correspond to semantically matched concepts from the literature.

**Table 2.**
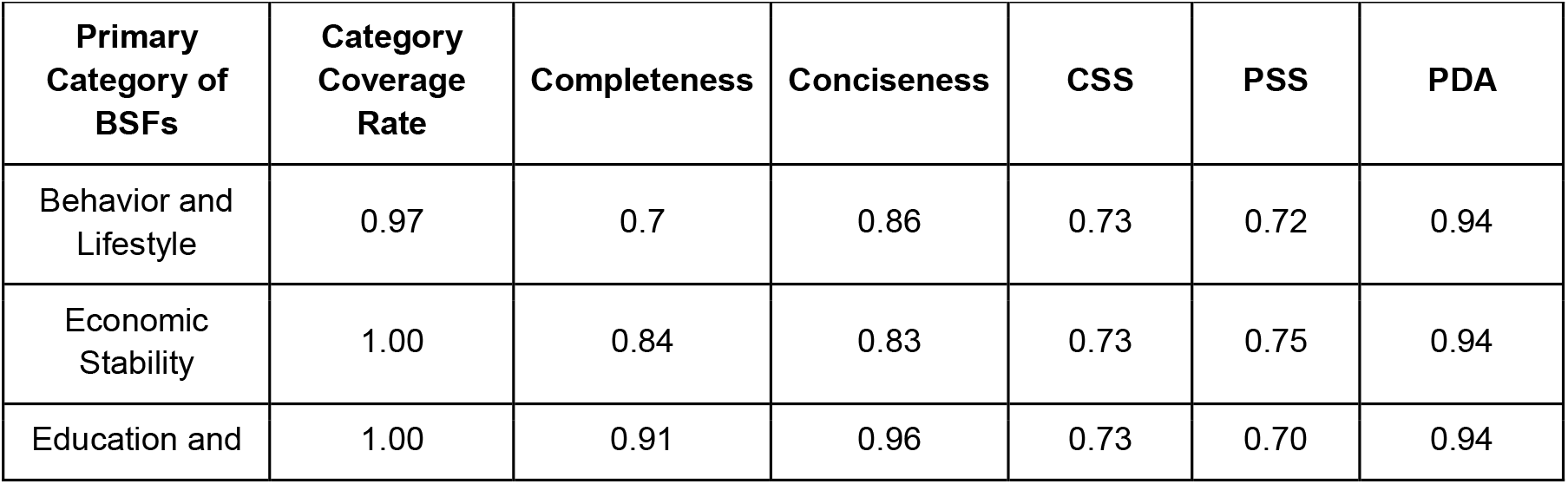
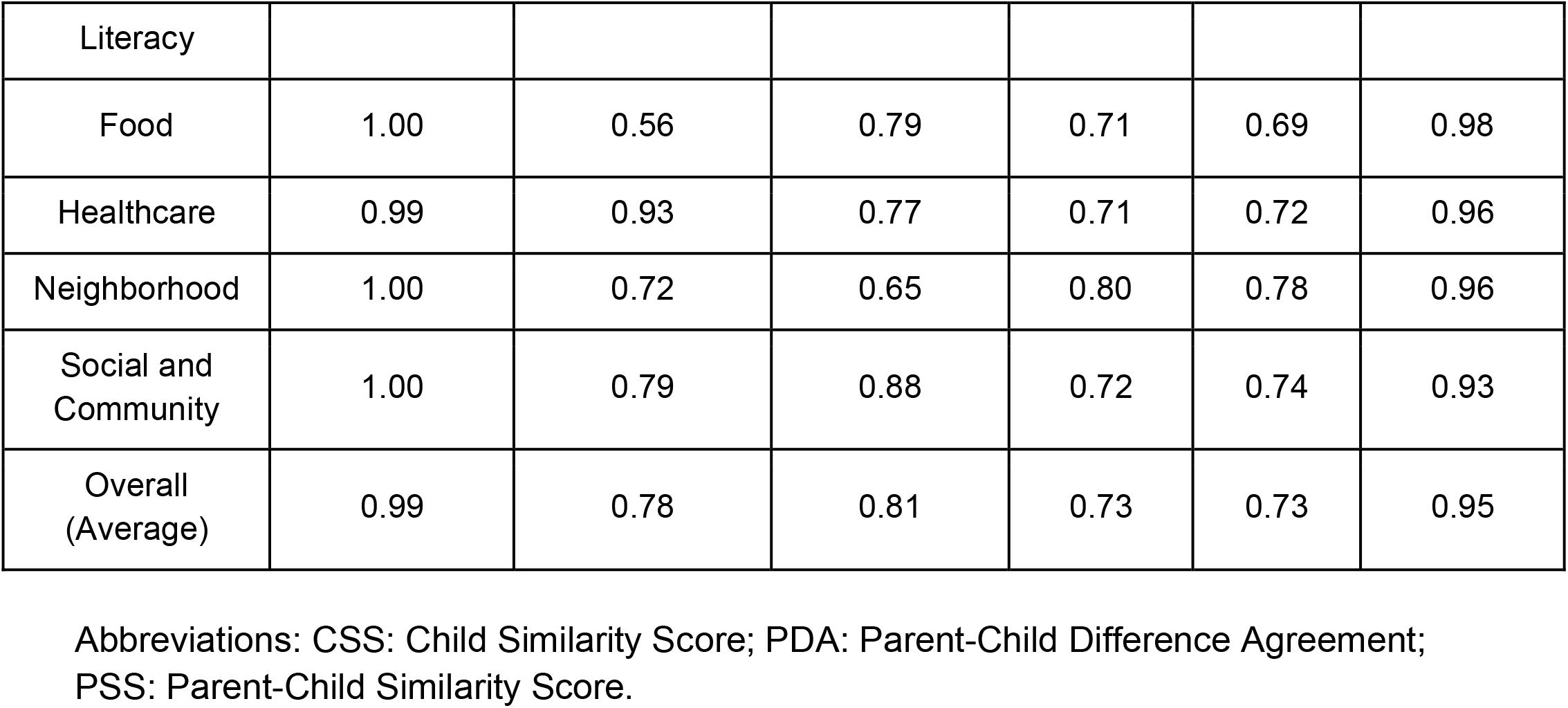
LLM-based automated evaluation results.

Semantic similarity metrics showed stable and coherent hierarchical organization across BSF categories, with CSS ranging from 0.71 to 0.80 and PSS ranging from 0.69 to 0.78. PDA was uniformly high (≥ 0.93), indicating consistent parent-child similarity patterns within concept families. Specifically, the Neighborhood category exhibited the strongest performance (CSS = 0.80, PSS = 0.78), reflecting a well-defined and semantically cohesive sub-ontology. In contrast, the Food category showed the lowest correctness scores (CSS = 0.71, PSS = 0.69), suggesting that its subclasses are semantically heterogeneous but equally distant from the parent concept. This pattern points to potential areas for structural enrichment, such as introducing intermediate classes to better distinguish parent-children classes. To demonstrate how correctness metrics can inform refinement, we examined the Food category using a CSS-PSS quadrant plot (Figure 5). Within the category, the Diet (CSS = 0.81, PSS = 0.63) appears in the second-quadrant region where its children cluster tightly, but the parent is overly broad, suggesting the need for an intermediate category. In contrast, “Food Insecurity” and “Lack of Adequate Food” demonstrate moderately high PSS but low CSS, indicating that these underspecified families require either additional subclasses or consolidation.

**Figure 5.**
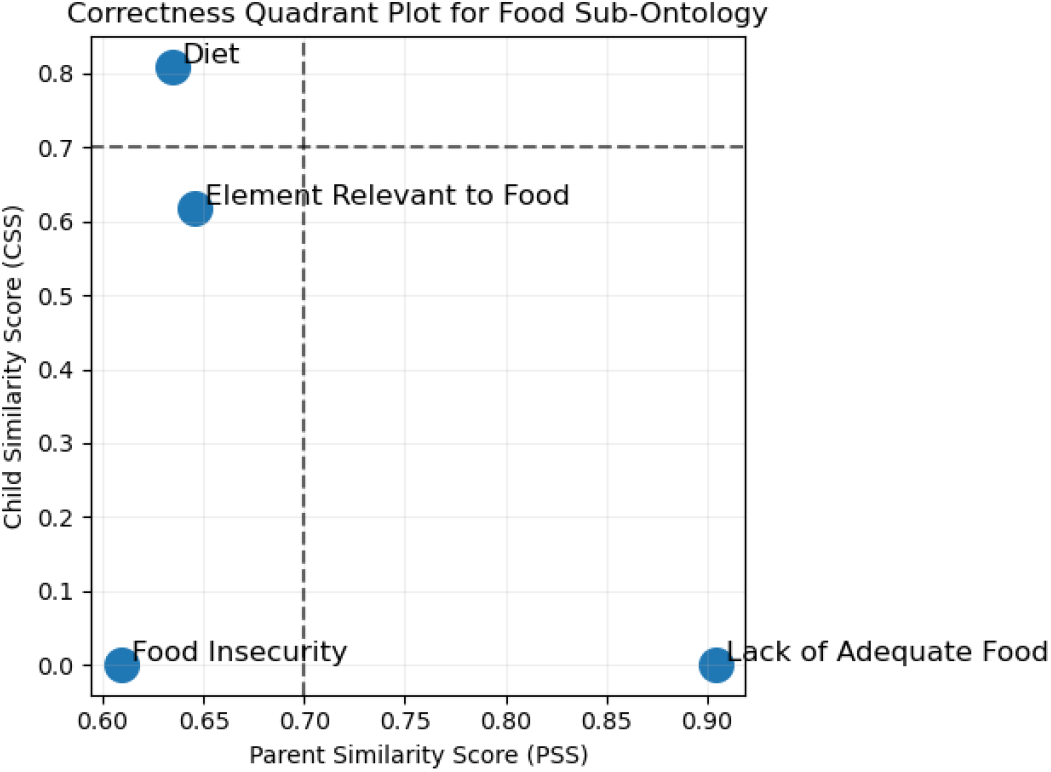
CSS-PSS quadrant plot for Food sub-category. Note: The x- and y-axis thresholds are set to 0.7. Abbreviations: CSS: Child Similarity Score; PSS: Parent-Child Similarity Score.

### Use Case

To demonstrate the representation capability of BSO-AD, we created a synthetic clinical note for an AD patient case (**Box 1**) using ChatGPT^42^. As illustrated in **Figure 6**, the patient is an elderly woman presenting with progressive memory loss over the past 18 months and depressive symptoms assessed by Geriatric Depression Scale, ultimately diagnosed with mild cognitive impairment due to AD. Beyond core clinical features, the case captures a rich set of BSFs, including widowhood following spouse death five years ago, living alone, social isolation, financial insecurity, lack of transportation for daily life, and a highly processed food diet. These factors are formally represented using BSO-AD classes and interconnected through object properties such as *hasAdverseLifeExperience, hasLivingStatus*, and *contributesTo*, enabling explicit modeling of pathways through which the BSFs influence AD onset and progression. Temporal attributes (e.g., duration of symptoms, timing of spouse death) and assessment instruments (e.g., Geriatric Depression Scale, Alzheimer’s Disease Assessment Scale) are also presented, demonstrating the ontology’s capacity to integrate longitudinal trajectories and evaluative dimensions. This use case illustrates how BSO-AD bridges clinical characteristics and BSFs, supporting high-resolution, multi-dimensional patient representation that can inform downstream tasks such as precise cohort discovery, causal inference, and longitudinal risk modeling.

**Figure 6.**
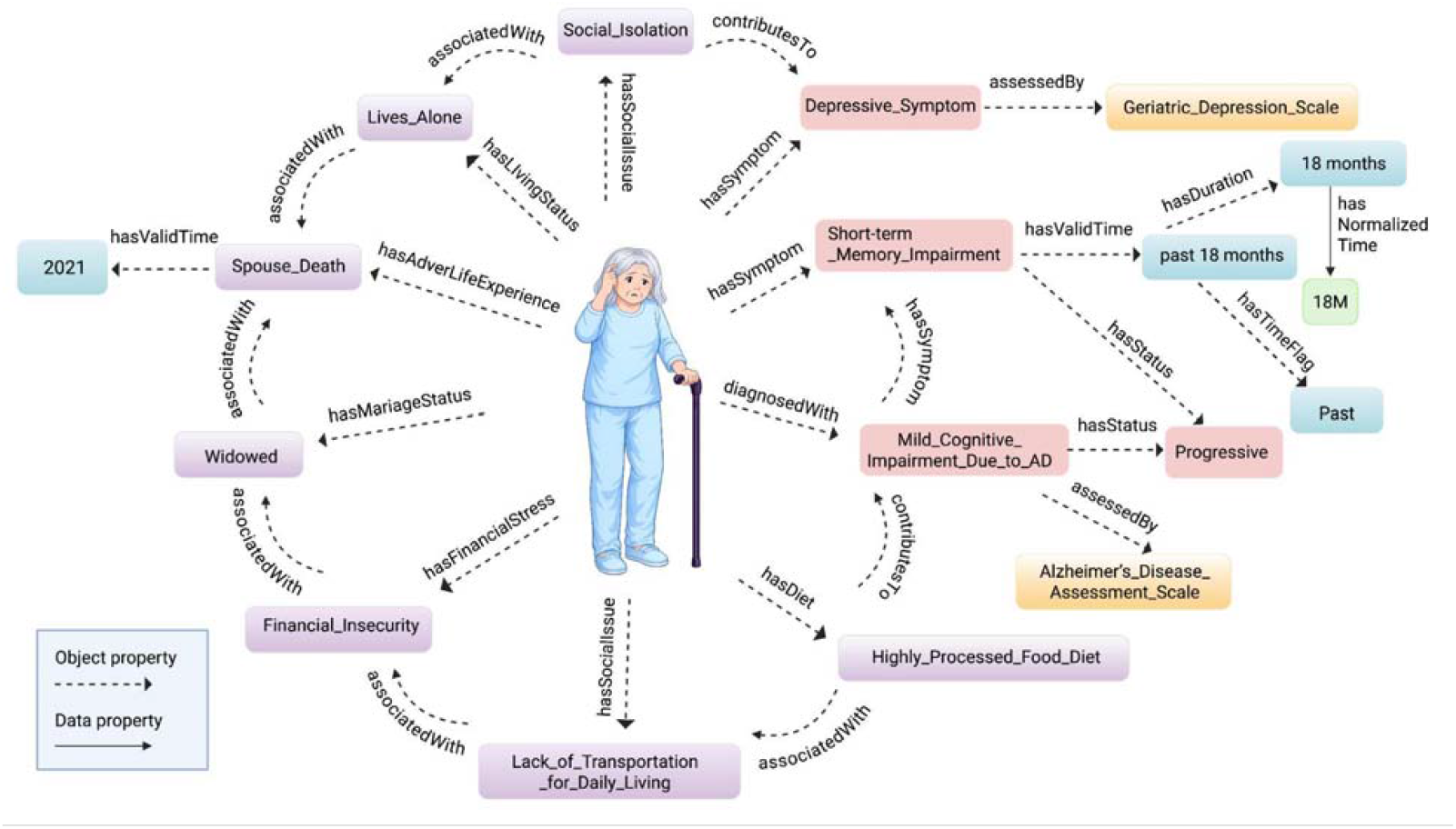
Knowledge representation of clinical, behavioral, and social factors in a synthetic AD patient case using BSO-AD

#### Box 1.

Synthetic Note*

The patient is a 76-year-old female with a prior diagnosis of mild cognitive impairment, now presenting with progressive short-term memory loss over the past 18 months. The patient lives alone following the death of her spouse in 2021. She endorses persistent feelings of loneliness and reduced participation in previously enjoyed social activities. She reports financial strain and limited access to transportation, which restricts her ability to attend medical appointments and obtain groceries. Her diet primarily consists of processed foods. The patient was assessed by the Geriatric Depression Scale for mild depressive symptoms. Alzheimer’s Disease Assessment Scale evaluation indicates progressive cognitive decline concerning early Alzheimer’s disease. Significant contributing social and behavioral factors include social isolation, poor diet, and possible depression.

*\*Note: This patient note is generated by ChatGPT. No real patient data was used, and no human subjects were involved. Thus, this section is exempt from the Health Insurance Portability and Accountability Act (HIPAA) considerations*.

## DISCUSSION

BSO-AD is the first specialized ontology designed to standa-termrdize and harmonize ADRD-related BSFs knowledge from heterogeneous sources. It provides a formally structured and semantic representation of BSFs tailored for ADRD by integrating functionality from existing ontologies, including SDoHO, DROADO, and TEO. In addition, BSO-AD incorporates standardized SDoH codes (ICD-10-CM Z55-Z65) and ADRD-related concepts from AD-Onto, ICD-9-CM, and ICD-10-CM. By reusing and extending existing ontologies, BSO-AD promotes interoperability and improves applicability in downstream tasks such as EHR-based knowledge integration.

BSO-AD introduces a multi-level hierarchy of semantic relations (object properties) that represent both functional and temporal associations between BSFs and ADRD (e.g., “increasesRiskOf” and “associatedWithShorterTime”). In addition to these direct relationships, BSO-AD captures indirect associations through intermediate biological entities, enabling representation of underlying biological mechanisms. These fine-grained relations support modeling how BSFs influence ADRD onset, progression, and outcomes across both behavioral-social and biological dimensions, facilitating the integration of multiscale real-world evidence to inform medical decision-making.

The evaluation results highlight several notable characteristics of BSO-AD. First, at the overall ontology level, the high scores of LLM-based category coverage and embedding-based completeness indicate that the ontology represents a stable set of concepts that recurs across both literature extractions and semantic similarity modeling. Despite the variability in terminology and granularity across the literature, concepts identified by LLMs consistently map to ontology classes in the embedding space. This alignment suggests that BSO-AD is robust to linguistic variation and captures the key constructs underlying BSF-ADRD research. Second, the correctness scores (CSS and PSS) fall in a moderate range, reflecting that BSF concepts are semantically related yet conceptually distinct. Unlike domains with rigid hierarchical distinctions (e.g., Anatomy or Disease), BSF constructs often exhibit overlapping conceptual boundaries. Related classes, such as “Social Isolation” and “Lives Alone” tend to share contextual usage while representing distinct aspects of a broader construct. Collectively, these findings suggest that BSO-AD provides a semantically coherent and structurally stable representation of BSFs, and is well-suited for downstream computational applications.

The evaluation results also inform directions for further ontology enhancement and enrichment. For instance, the Healthcare domain shows high completeness but low conciseness, indicating that some classes are underrepresented in the ADRD-focused socio-behavioral literature used for evaluation. This may reflect limitations in the literature query scope or indicate that these concepts can be better captured in alternative data sources such as EHRs. In addition, the quadrant plot (**Figure 5**) highlights areas within the Food hierarchy that exhibit insufficient granularity or have overly heterogeneous groupings. This analytic approach can be applied to systematically identify and prioritize targets for ontology refinement.

Looking ahead, we plan to enrich BSO-AD through large-scale literature- and EHR-based data mining, as well as incorporating elements from additional ontologies and standards, such as the ICD-11 “Factors influencing health status” category. Compared with ICD-10-CM, ICD-11 adopts a more interoperable approach, with enhanced capabilities in integrating with modern digital health systems and supporting global data exchange^43^. Incorporating ICD-11 SDoH codes will enhance the breadth and depth of the ontology, enabling more precise representation and harmonization of BSF data. In parallel, we will refine the LLM-driven evaluation pipeline by incorporating human-in-the-loop review, human-annotated gold standard corpora, and advanced proprietary LLMs such as OpenAI’s GPT series and Google’s Gemini, to boost its accuracy, effectiveness, and generalizability. Furthermore, we will explore integrating BSO-AD as a structured knowledge foundation to augment LLM-based frameworks, supporting more robust entity extraction, normalization, and grounded reasoning in ADRD-related BSF analysis. We will also utilize BSO-AD as a semantic backbone to guide the development and refinement of large-scale knowledge graphs, facilitating evidence-based hypothesis generation and knowledge discovery.

## CONCLUSION

The BSO-AD establishes a unified, semantically rich knowledge representation of BSFs and ADRD concepts and their interrelationships. By integrating standardized vocabularies, curated relationships, and an LLM-assisted evaluation pipeline, BSO-AD lays a robust foundation for scalable harmonization and analysis of BFSs within the context of ADRD, potentially supporting advanced computational modeling and knowledge discovery across interdisciplinary studies.

## SUPPLEMENTARY MATERIAL

Supplementary 1.csv

Supplementary 2.docx

Supplementary 3.csv

## DATA AVAILABILITY

The BSO-AD is publicly available through GitHub at https://github.com/Tao-AI-group/BSO-AD and through Zenodo at https://doi.org/10.5281/zenodo.20328263.

## CODE AVAILABILITY

The source code used for ontology development and LLM-assisted ontology evaluation is publicly available at: https://github.com/Tao-AI-group/BSO-AD.

## AUTHOR INFORMATION

### Authors and Affiliations

**Department of Artificial Intelligence and Informatics, Mayo Clinic, Jacksonville, FL, USA**

Haifang Li, Yue Yu, Avanti Bhandarkar, Rakesh Kumar, Isaac H. Clark, Weiguo Cao, Fang Li & Cui Tao

**Department of Computer Science, Emory University, Atlanta, GA, USA**

Yutong Hu

**Department of Neuroscience, Mayo Clinic, Jacksonville, FL, USA**

Na Zhao

### Contributions

CT conceptualized the study, secured funding and resource support, supervised the overall project, and critically revised the manuscript. FL co-supervised the study and critically revised the manuscript. HL, YY, and AB drafted the manuscript. HL constructed the ontology, extracted object properties from the literature, and revised the manuscript. YY developed the ICD code mappings. RK enriched the ADRD-related concepts. AB developed the LLM-based ontology evaluation pipeline. RK and IC conducted the ontology manual review. WG and YH provided technical support. NZ guided BSF-ADRD relationship design. All authors reviewed and approved the final version of the manuscript.

## ETHICS DECLARATIONS

### Competing Interests

The authors declare no competing interests.

## FUNDING

This study was supported by the National Institute of Aging under U01AG088076, R01AG072799, and U24AG088019, and the National Institute of Mental Health under U24MH136069.

## Footnotes

1 MedGemma was selected based on its superior performance in preliminary evaluations of medical entity recognition relative to the other evaluated models.

## Notes

### Competing Interest Statement

The authors have declared no competing interest.

### Summary of Updates

This revised version includes updates to the ontology resource, GitHub and Zenodo links.

