## Supplementary 2 for "BSO-AD: An Ontology for Representing and Harmonizing Behavioral Social Knowledge in ADRD" (from another version of this preprint: version 1, 2026-03-31)

### Supplement 2

#### 1. Supplementary Tables

**Table S1.** ADRD-related concepts from ICD-9-CM and ICD-10-CM

| ADRD-related Concepts | ICD Codes | |
|---|---|---|
| | ICD-9-CM | ICD-10-CM |
| Dementia | 290.*, 797.0, 331.1, 331.82 | F01.5, F02.8, F03.9, F04, F05, F06.0, F06.1, F06.2, F06.3, R41.81, G31.83, G31.0 |
| Alzheimer's Disease | 331.0 | G30.* |

*represents including all subclasses

**Table S2.** PubMed search query for retrieving BSF-ADRD literature for relationship construction: (queried on 04/28/2025)

| |
|---|
| ((Alzheimer Disease[Title/Abstract]) OR (Dementia[Title/Abstract]) OR (Dementia[MeSH Terms]) OR (Alzheimer Disease[MeSH Terms]))<br>AND<br>((Social Determinants of Health[Title/Abstract]) OR (Social Determinants of Health[MeSH Terms])) |

**Table S3.** Object properties (indirect associations) between BSFs and ADRD, as well as their intermediate biological nodes

| Domain | Range | Object Properties |
|---|---|---|
| ADRD | BSF | associatedWith |
| ADRD | gene | regulates/associatedWith |
| ADRD | pathology | manifestsAs/associatedWith |
| ADRD | pathway | regulates/associatedWith |
| BSF | ADRD | increasesRiskOf/decreasesRiskOf* |
| BSF | gene | modulates/activates/suppresses |

| | | |
|---|---|---|
| BSF | pathology | increases/decreases |
| BSF | pathway | modulates/dysregulates |
| gene | ADRD | increasesRiskOf/decreasesRiskOf/causes |
| gene | BSF | associatedWith |
| gene | pathology | causes/increases/decreases |
| gene | pathway | participatesIn/regulates |
| pathology | ADRD | associatedWith |
| pathology | BSF | associatedWith |
| pathology | gene | activates/suppresses/associatedWith |
| pathology | pathway | perturbs/dysregulates |
| pathway | BSF | associatedWith |
| pathway | ADRD | causes/regulates/associatedWith |
| pathway | gene | activates/suppresses/regulates/associatedWith |
| pathway | pathology | causes/involvedIn/affectedBy |

Abbreviation: ADRD, Alzheimer's disease and related dementias; BSF, behavioral social factor

**Table S4.** Synonyms* of object properties (direct effect) of BSFs-ADRD

| **Object properties** | **Synonyms** |
|---|---|
| associatedWith | relatedTo, correlatesOf, correlatesWith, linkedTo, linkedWith |
| affects | impacts, influences |
| benefits | enhances, improves, promotes, supports |
| affectsRiskOf | correlatesWithRiskOf, accountForRiskOf, associatedWithRiskOf, attributesToRiskOf, contributesToRiskOf, correlatesRiskOf, impactsRiskOf, influencesRiskOf, aRiskOf, aRiskFactorFor, linkedToRiskOf, linkedWithRiskOf, predictsRiskOf, relatedToRiskOf |
| decreasesRiskOf | diminishesRiskOf, hasLowerOddsOf, lessensRiskOf, lowersRiskOf, mediatesRiskOf, mitigatesRiskOf, |

| | |
|---|---|
| | reducesRiskOf |
| increasesRiskOf | amplifiesRiskOf, escalatesRiskOf, exacerbatesRiskOf, increasesRiskFor, associatedWithIncreasedRiskOf, LinkedToIncreasedOddsOf, leadsToHigherRiskOf, raisesRiskOf |
| mitigates | alleviates |
| causes | aDriverOf |
| contributesTo | aContributorTo |
| hasHigherIncidenceOf | hasGreaterIncidenceOf |

*These synonyms were extracted from the PubMed literature.

**Table S5.** List of Abbreviations

| Abbreviations | Full Name |
|---|---|
| AD | Alzheimer’s Disease |
| ADMO | Alzheimer's Disease Map Ontology |
| ADNI | Alzheimer's Disease Neuroimaging Initiative |
| ADO | Alzheimer’s Disease Ontology |
| ADRD | Alzheimer’s Disease and Related Dementias |
| BSFs | Behavioral and Social Factors |
| BSO-AD | Behavioral Social Data and Knowledge Ontology for ADRD |
| CSS | Child Similarity Score |
| CUIs | Concept Unique Identifiers |
| DROADO | Drug Repurposing-Oriented Alzheimer’s Disease Ontology |
| EHRs | Electronic health records |
| FAIR | Findable, Accessible, Interoperable, and Reusable |
| LLM | Large Language Model |
| MeSH | Medical Subject Headings |
| NER | Named Entity Recognition |

| | |
|---|---|
| NLP | Natural Language Processing |
| OBO | Open Biological and Biomedical Ontologies |
| OMRSE | Ontology of Medically Related Social Entities |
| OWL | Web Ontology Language |
| PACO | Physical Activity Ontology |
| PDA | Parent Difference Agreement |
| PSS | Parent Similarity Score |
| SDoH | Social Determinants of Health |
| SDoHO | Social Determinants of Health Ontology |
| SMASH | Semantic Mining of Activity, Social, and Health Data |
| TEO | Time Event Ontology |
| UMLS | Unified Medical Language System |

### 2. Supplementary Texts

#### 2.1 Literature query and screening

##### 2.1.1 MeSH and Keyword-based Query for PubMed Literature Retrieval

((Alzheimer Disease[MeSH Major Topic]) OR (Primary Progressive Aphasia[MeSH Major Topic]) OR (Primary Progressive Nonfluent Aphasia[MeSH Major Topic]) OR (Vascular Dementia[MeSH Major Topic]) OR (CADASIL[MeSH Major Topic]) OR (Multi-Infarct Dementia[MeSH Major Topic]) OR (Frontotemporal Lobar Degeneration[MeSH Major Topic]) OR (Frontotemporal Dementia[MeSH Major Topic]) OR (Primary Progressive Nonfluent Aphasia[MeSH Major Topic]) OR (Huntington Disease[MeSH Major Topic]) OR (Lewy Body Disease[MeSH Major Topic]) OR (Mixed Dementias[MeSH Major Topic]) OR (Alzheimer[Title/Abstract])) **AND** ((Social Determinants of Health[MeSH Major Topic]) OR (Social Determinants of Health[Title/Abstract]) OR (sdoh[Title/Abstract]) OR (smoke[Title/Abstract]) OR (alcohol consumption[Title/Abstract]) OR (education[Title/Abstract]) OR (financial[Title/Abstract]) OR (behavior[Title/Abstract]))

16,241 - overall

10 years (2015-2025), 9,629 results (07/27/2025)
Human + English + Abstract not null, 6,106 results (07/28/2025)

**2.1.2 Prompt for literature screening**

Instructions:
Your task is to evaluate each candidate paper (based on the given title and abstract) against the following three criteria:
1. AD/ADRD Focus: The study must focus on Alzheimer's Disease or related dementias (AD/ADRD), including but not limited to: Alzheimer Disease, Primary Progressive Aphasia, Primary Progressive Nonfluent Aphasia, Vascular Dementia, CADASIL, Multi-Infarct Dementia, Frontotemporal Lobar Degeneration, Frontotemporal Dementia, Primary Progressive Nonfluent Aphasia, Huntington Disease, Lewy Body Disease, Mixed Dementias.
2. Behavioral and Social Factors Focus: The study must explore the relationship between AD/ADRD and behavioral or social factors (e.g., behavioral symptoms, social determinants of health, interpersonal relationships, caregiver burden).
3. Human study: The study must involve human participants or data. Exclude studies based exclusively on animal models.

Output:
For each criterion, return only 'true' or 'false' for each of the criteria. Next, provide a final decision ('include' or 'exclude') along with a brief explanation. Make your assessments solely based on the information contained in the title and abstract.
Use the exact output format below:

```
{
    "paper_id": "<paper ID>",
    "ADRD_focus": true/false,
    "behavior_social_focus": true/false,
    "human_study": true/false,
    "final_decision": {
        "include": true/false,
        "reason": "..."
        }
}
```

Input:
Paper ID: {paper_id}

Title: {title}
Abstract: {abstract}

**Selected articles: 2119 (08/06/2025)**

#### 2.2. LLM-based Concept Extraction and Ontology Coverage Evaluation

##### 2.2.1 Prompt 1

Requirements:
Your task is to extract ADRD-related {factor} entities (single or multiple word phrases) and associated sentences from PubMed abstracts.

Format:
- Factor Relevance: Classify whether the given PubMed abstract represents a study on the impact of {factor} on ADRD
- Next, identify potential entities for:
i. {factor} factors and
ii. AD/ADRD
- For each extracted entity:
i. Provide the source sentence that shows the association between {factor} and ADRD
ii. Give an explanation within 10 words justifying the relevance of the entity.

#### Quality:
- While assessing factor relevance, ensure that the extracted entities are NOT related to symptoms, medications, genes, etc. related to ADRD when factor_relevance is No.
- Solely focus on the documented impact of the BSSR factor: {factor} on ADRD
- Only extract entities associated with ADRD. Ignore sentences that read like background or general related information.
- DO NOT hallucinate or change the names of entities or categories, as well as the source sentence.
- Make your assessments solely based on the information contained in the given Data (title and abstract).

Output:
Here is an example of a template for the output. DO NOT return any other text other than the output json.
```json
```

```
{
"paper_PMID": "<PMID>",
"factor_relevance":<whether relevant to {factor}--Yes/No>
"potential_entities": [{
"entity_name": <the entity name>,
"source_sentence": <sentence evidence from the PubMed abstract>,
"category": <assigned category -- ADRD/{factor}>,
"explanation":<justification for annotation>},
...
{
...
}
]
}
```

Input:
Paper ID: {paper_id}
Title: {title}
Abstract: {abstract}

##### 2.2.2 Prompt 2

- System Instruction:
    - Context:

You are an expert in Behavioral and Social Science Research (BSSR), which involves the systematic study of how behaviors and social factors influence health. Currently, you are focusing on investigating the documented impact of {factor} factors on Alzheimer's disease and related dementias (ADRD).

- Data

As input, you will be given the following:

- PubMed title
- PubMed abstract
- {factor} related entity
- source sentence which contains the entity

- Task:

Your task is to classify the ADRD-related {factor} entity (single or multi-word phrases) using the associated sentence into one of the following classes based on the provided definitions: {class_list+Undefined}

- **Main Prompt**

**Format**:
- Factor Relevance: Verify whether the given entity truly represents {factor} factor
- Factor class: Classify the entity in the source sentence in one or more of the given classes. If the entity cannot be classified into any of the classes, classify it as "Undefined"
- Explanation: Give an explanation within 10 words justifying the relevance of the entity to the assigned class.

**Quality**:
- While verifying factor relevance, ensure that the extracted entities are NOT related to symptoms, medications, genes, etc., related to ADRD.
- Solely focus on the documented impact of the BSSR factor: {factor} on ADRD
- Only extract entities associated with ADRD. Ignore sentences that read like background or general related information.
- DO NOT hallucinate or change the names of entities or categories, as well as the source sentence.
- Make your assessments solely based on the information contained in the given Data.

**Examples**:
Below are a few examples of Factor class classification.
Note that if an entity/sentence can be assigned to a factor class, then the factor relevance must be Yes.

{examples}

**Output**:
Here is an example of a template for the output. DO NOT return any other text other than the output json.
```json
    {
    "paper_PMID": "<PMID>",
    "entity_name": "<entity name>",
    "source_sentence": "<sentence evidence>",
    "factor_relevance":"<Yes/No>"
    "factor_class": ["<factor class label 1>", "<factor class label 2>"],
    "explanation":"<justification for annotation>",
    }
```

**Input:**

Paper ID: {paper_id}
Title: {title}
Abstract: {abstract}
Entity name: {entity_name}
Source sentence: {source_sentence}

#### 2.3. Ontology evaluation metrics

The following metrics are reproduced from [39].

**Child Similarity Score (CSS):** the mean cosine similarity between every pair of siblings in a Concept Family (CF)

CSS (CF) = $\frac{1}{M} \sum_{i=1, j=i+1}^{M-1}$ *similarity* ($C_i$,$C_j$)

Where,
M is the number of child concepts in a CF, Ci and Cj are sibling (child) concepts

**Parent-Child Similarity Score (PSS):**

PSS (CF) = $\frac{1}{M} \sum_{i=1}^{M}$ *similarity* ($C_i$,$C_p$)

Where,
Cp is the parent concept and
M is the number of child concepts

**Parent-Child Difference Agreement (PDA):**

$$PDA\ (CF) = 1 - \sqrt{\frac{1}{M-1} \sum_{i=1}^{M} [Similarity\ (Ci, Cp)\ -\ PSS\ (CF)]^2}$$

#### 2.4 Detailed results for category coverage evaluation

**<u>Step 1:</u> Entity Extraction and Binary Classification**

| **BSF Primary Category** | **AD/ADRD** | **BSF** | **Total** |
|---|---|---|---|
| **Behavior and Lifestyle** | 2361 | 3264 | 5625 |
| **Economic stability** | 811 | 847 | 1658 |

| | | | |
|---|---|---|---|
| **Education and Literacy** | 1877 | 1403 | 3280 |
| **Food** | 439 | 450 | 889 |
| **Healthcare** | 1030 | 1789 | 2819 |
| **Neighborhood** | 281 | 287 | 568 |
| **Social and Community** | 2420 | 3389 | 5809 |

**<u>Step 2</u>: BSF (single factor) relevance detection (binary) and Ontology Level 3 classification (multi-label)**

**<u>Factor: Behavior and Lifestyle</u>**

| **Factor subclasses** | **#entities** |
|---|---|
| Total relevant | 2702 |
| Undefined | 77 |
| Behavior change | 1381 |
| diet | 326 |
| Physical activity | 479 |
| Sexual behavior | 5 |
| Sleep | 351 |
| Spirituality and religion | 33 |
| Substance use | 388 |
| Treatment adherence and compliance | 11 |

**<u>Factor: Economic stability</u>**

| **Factor subclasses** | **#entities** |
|---|---|
| Total relevant | 778 |
| Undefined | 0 |
| Socioeconomic status | 339 |
| Financial stress | 188 |

| | |
|---|---|
| Income | 111 |
| Expense | 109 |
| Healthcare payment source | 56 |
| Employment | 53 |
| Problem related to housing and economic circumstance | 49 |
| Wealth | 33 |

**Factor: Education and Literacy**

| **Factor subclasses** | **#entities** |
|---|---|
| Total relevant | 1157 |
| Undefined | 0 |
| Education level | 820 |
| Years of education | 308 |
| Literacy | 107 |
| Education quality | 83 |
| Access to education | 27 |
| Problem related to education and literacy | 6 |
| Early childhood education | 4 |

**Factor: Food**

| **Factor subclasses** | **#entities** |
|---|---|
| Total relevant | 329 |
| Undefined | 0 |
| Diet | 322 |
| Food insecurity | 4 |
| Access to health and nutritious options | 3 |

**Factor: Healthcare**

| **Factor subclasses** | **#entities** |
|---|---|
| Total relevant | 1523 |

| | |
|---|---|
| Undefined | 2 |
| Health access | 296 |
| Quality of care | 551 |
| Treatment and medication | 324 |
| Healthcare visit | 272 |
| Healthcare payment source | 128 |

**Factor: Neighborhood**

| **Factor subclasses** | **#entities** |
|---|---|
| Total relevant | 266 |
| Undefined | 0 |
| Socioenvironmental neighborhood | 172 |
| Geographic location | 79 |
| Physical environment | 25 |
| Problems related to physical environment | 4 |

**Factor: Social and Community**

| **Factor subclasses** | **#entities** |
|---|---|
| Total relevant | 3052 |
| Undefined | 0 |
| Social environment | 1006 |
| Social support | 879 |
| Material deprivation | 396 |
| Social norms and attitude | 288 |
| Culture | 231 |
| Social activity | 199 |
| Community engagement | 150 |
| Social isolation | 137 |
| Problems related to social environment | 106 |
| Incarceration | 11 |

| | |
|---|---|
| Safety concern | 6 |
| Psychosocial factor | 523 |
| Social deprivation | 0 |
